# Impact of COVID-19 on the outreach strategy of cancer social service agencies in Singapore: A pre-post analysis with Facebook data

**DOI:** 10.1101/2020.11.11.20229740

**Authors:** Kieran Ethan Tan, Aravind Sesagiri Raamkumar, Hwee Lin Wee

## Abstract

The Singapore government implemented multiple restrictive measures as the novel coronavirus infection (COVID-19) spread through the community, thereby affecting the support service of cancer-related social service agencies (cancer-SSAs). We are interested to understand how Singapore’s cancer-SSAs utilized the social media platform Facebook to overcome the restrictions which were introduced due to COVID-19. Facebook posts from cancer-SSAs 365 Cancer Prevention Society (365CPS) and Singapore Cancer Society (SCS) between comparable periods in 2019 and 2020 were extracted. These posts were categorized using a classification scheme specifically developed for this study. Statistical analyses were performed to determine if there was a significant difference in the frequency of posts between 2019 and 2020, and across three specific periods in 2020. Results indicate that 365CPS appears to have adapted to the pandemic by increasing their posting frequency on Facebook in 2020, but the same was not evident for SCS. However, both SSAs tweaked their social media outreach strategy in line with social distancing measures, publishing posts detailing activities that beneficiaries can participate from home such as healthy recipes and virtual events. SSAs can scale up their efforts to achieve a higher level of health promotion and support for their beneficiaries.

## 1. Introduction

The novel coronavirus (COVID-19) was discovered in Wuhan, China on December 31^st^, 2019 [1]. Infected patients show mild to moderate respiratory symptoms, and the disease is transmitted via respiratory droplets or saliva when one sneezes or coughs [2]. Most people can recover on their own, but others are at a higher risk of developing severe complications when infected, such as those with underlying medical conditions such as cancer [3]. On March 11^th^ 2020, World Health Organization (WHO) recognized the virus as a pandemic [4].

The first person infected with COVID-19 in Singapore on January 23^rd^, 2020 marked the start of Singapore’s fight against the pandemic [5]. The government updated the Disease Outbreak Response System Condition (DORSCON) status to Orange on February 7^th^ 2020, and introduced new measures to minimize transmission in the community [6]. The circuit breaker (CB) i.e. lockdown, was announced on April 7^th^ 2020 following increased cases in the community [7]. Subsequently, increased restrictions were implemented to curb the transmission of the diseases. Services such as rehabilitation had to be suspended. Singapore entered Phase 1 of reopening on June 1^st^ 2020, gradually easing restrictions on previously restricted services. However, social distancing measures are still being implemented to curb spread of the virus [8].

Social service agencies (SSA) provide aids to their beneficiaries. These aids include rehabilitation facilities that provide counselling services for patients. However, the COVID-19 pandemic has affected how they provided support whilst adhering to social distancing guidelines [9–11]. For example, Singapore Cancer Society (SCS), a cancer-related SSA (cancer-SSA) specializing in providing support to cancer patients, had many of their rehabilitation and transport services suspended or restricted. As communication with beneficiaries became limited due to these restrictions, the level of support these SSAs could provide during the COVID-19 pandemic was severely diminished.

A total of 71,265 cancer cases were reported between 2013-2017 in Singapore [12], with roughly 39 people diagnosed on a daily basis [13]. This number is still increasing, and with it, the need to support cancer patients increases. Support becomes crucial during the pandemic, as cancer patients cannot receive the same level of support compared to the pre-COVID-19 period. Cancer patients may be at a higher risk of contracting the coronavirus during this time, and they may not want to leave their home to attend essential cancer care sessions. The prolonged stay at home also prevents them from receiving face-to-face counselling from SSAs, which may lead to detrimental consequences to their mental health and wellbeing. Since cancer-SSAs are unable to continue with existing services as per normal, they will have to develop new strategies to communicate and guide beneficiaries in taking ownership of their health at home.

Many businesses have turned to social media as a platform to connect with consumers since in-person communication has become limited during the pandemic. A global study showed that media consumption increased by 61% compared to normal period, with an 40% increase in social media consumption on Facebook from the start of the pandemic to April 2020 [14]. Overall, social media presence for businesses is important during this period for communicating with their consumers. This statement applies to cancer-SSAs as well. Prior to the pandemic, these SSAs already had an online presence to connect and communicate with their beneficiaries. This pandemic has turned social media into a key platform for cancer-SSAs to share updates and information with cancer patients, but it remains to be seen whether these SSAs have improved their social media presence during the pandemic.

A previous study evaluated the role that social media played in health promotion in Singapore [15]. The study focused on Facebook posts published by the Singapore Health Promotion Board (HPB), analyzing the contents of the posts, and quantifying the level of audience engagement. They found that Facebook posts were well structured to effectively promote health-related information and activities while being warmly received by online audiences. Other studies have concluded that SSAs benefit from social media in improving current communication channels [16] and there is potential in social media for boosting health promotion opportunities, and behavioral changes for their audience [17].

However, no studies have been conducted to assess the effectiveness of social media for cancer-SSAs in Singapore in sharing information and promoting heath to their followers. Furthermore, no studies have been conducted to evaluate how the COVID-19 pandemic has affected the support activities of cancer-SSAs and their social presence. This study aims to conduct a social media analysis to analyze the trends of cancer-SSA’s Facebook posts during the COVID-19 pandemic. The first objective of the study is to compare the number of posts posted by each cancer-SSA between 2019 and 2020, and across three nominated periods in 2020. The second objective is to evaluate the content of these posts to ascertain whether the COVID-19 pandemic has had any impact in the types of posts shared by the SSAs. Two cancer-SSAs were identified for the purposes of this study. They are Singapore Cancer Society (SCS) and 365 Cancer Prevention Society (365CPS).

## 2. Data and Methods

### 2.1 Data Extraction and Timeline

We identified Facebook posts by cancer-SSAs in Singapore to be the unit of analysis for the study. Data was extracted from Facebook using the Facepager software [18,19]. The timeframes for data collection were (i): January 23^rd^ 2019 to June 1^st^ 2019, and (ii) January 23^rd^ 2020 to June 1^st^ 2020. January 23^rd^ was selected as the starting date as it was the same date in 2020 when the first person in Singapore was infected with COVID-19. June 1^st^ was selected as the ending date as it was the start of Phase 1 of post-CB in 2020 when more restrictions had been lifted. For the purpose of comparison, corresponding timeframes were selected in 2019.

2020 was split further into the following three periods to reflect the evolution of policy measures pertaining to COVID-19: (i) Period 1 (P1): January 23^rd^ 2020 to February 7^th^ 2020, (ii) Period 2 (P2): February 8^th^ 2020 to April 7^th^ 2020 and (iii) Period 3 (P3): April 8^th^ 2020 to June 1^st^ 2020. Period 1 (P1) represented the period from the start of COVID-19 in Singapore until the implementation of DORSCON Orange in Singapore. Period 2 (P2) represented the period from the start of DORSCON Orange in Singapore until the start of CB. Period 3 (P3) represented the period from the start of CB until the end of CB. Singapore’s Phase 1 post-CB was not included in this study as we wished to look at how the restrictive nature of CB affected the ways in which cancer-SSAs provided support and services to beneficiaries online. Social distancing measures and restrictions varied among the three periods, and different restrictions might have caused cancer-SSAs to adopt different strategies during each segment.

### 2.2 Exclusion Criteria

We did not include posts published on other social media platforms by the two cancer-SSAs. Other cancer-related societies such as Children’s Cancer Foundation and Breast Cancer Foundation were too specific in terms of the type of cancer or the target demographic. Hence, they were not included in this study. Posts without any text content were not analyzed in this study. Photos and videos were not included in the analysis. Non-English posts were not considered for the study.

### 2.3 Developing the Classification Categories for the Facebook Posts

Two coders classified the Facebook posts into their respective categories. Intercoder reliability was calculated based on the method described in Wimmer and Dominick [20]. Coders were briefed on the coding process and the classification scheme. A one-hour training session was conducted for the coders to familiarize themselves with the scheme and procedure using posts published by the two SSAs outside of the study timeframe, as described by Lacy et al [21]. The first pilot study was conducted using 50 posts from the study. Krippendorf’s alpha coefficient was calculated and used to assess intercoder reliability, with a coefficient of 0.80 or higher being deemed acceptable. Results from the first pilot study was sub-optimal and a second pilot study using 50 different posts from the study was conducted. A coefficient greater than 0.80 was achieved for the second pilot study. Subsequently, coding began for all posts from the study, with the primary coder coding for all Facebook posts, and the secondary coder coding 104 out of 244 posts to ensure the categorization of these Facebook posts remain reliable. The number of posts calculated for each pilot study and the actual study for reliability testing was based on the equation suggested by Lacy and Riffe for nominal content categories [22].

### 2.4 Statistical Analysis

For comparing the number of Facebook posts across time periods, Poisson regression was used. To compare the proportion of posts assigned to a particular category between the time periods for each category, the Chi-square test and Fisher’s exact test were used. All statistical tests were two-tailed and were performed at a significance level of 0.05. Two separate comparisons were tested: the first comparison between each year, and the second comparison between each nominated period in 2020. 2019 was used as a baseline to see how COVID-19 affected the cancer-SSA’s social media presence in 2020. The comparison between each nominated period in 2020 was conducted to evaluate if the different levels of social distancing measures adopted in each period has had a significant impact in the number and types of posts being published by the cancer-SSAs.

## 3. Results

### 3.1 Posting Frequency of Cancer-SSAs in Facebook

Figure 1 illustrates the number of monthly posts published by 365CPS and SCS for the analysis periods in 2019 and 2020. A total of 97 posts were extracted from 365CPS’s Facebook page. 96 posts were included in the study, with 1 post being excluded as its message was in Chinese. For all six months, the number of posts was higher in 2020 (n=66) compared to 2019 (n=30) (z=3.58, p-value<0.001). Additionally, the number of posts in 2020 were different across the three nominated periods at a statistically significant level (z=2.39, p-value=0.0171). 365CPS began publishing more posts since February 2020, coinciding with the announcement of DORSCON Orange in Singapore, with March 2020 recording the highest number of monthly posts (n=16).

**Fig.1.**
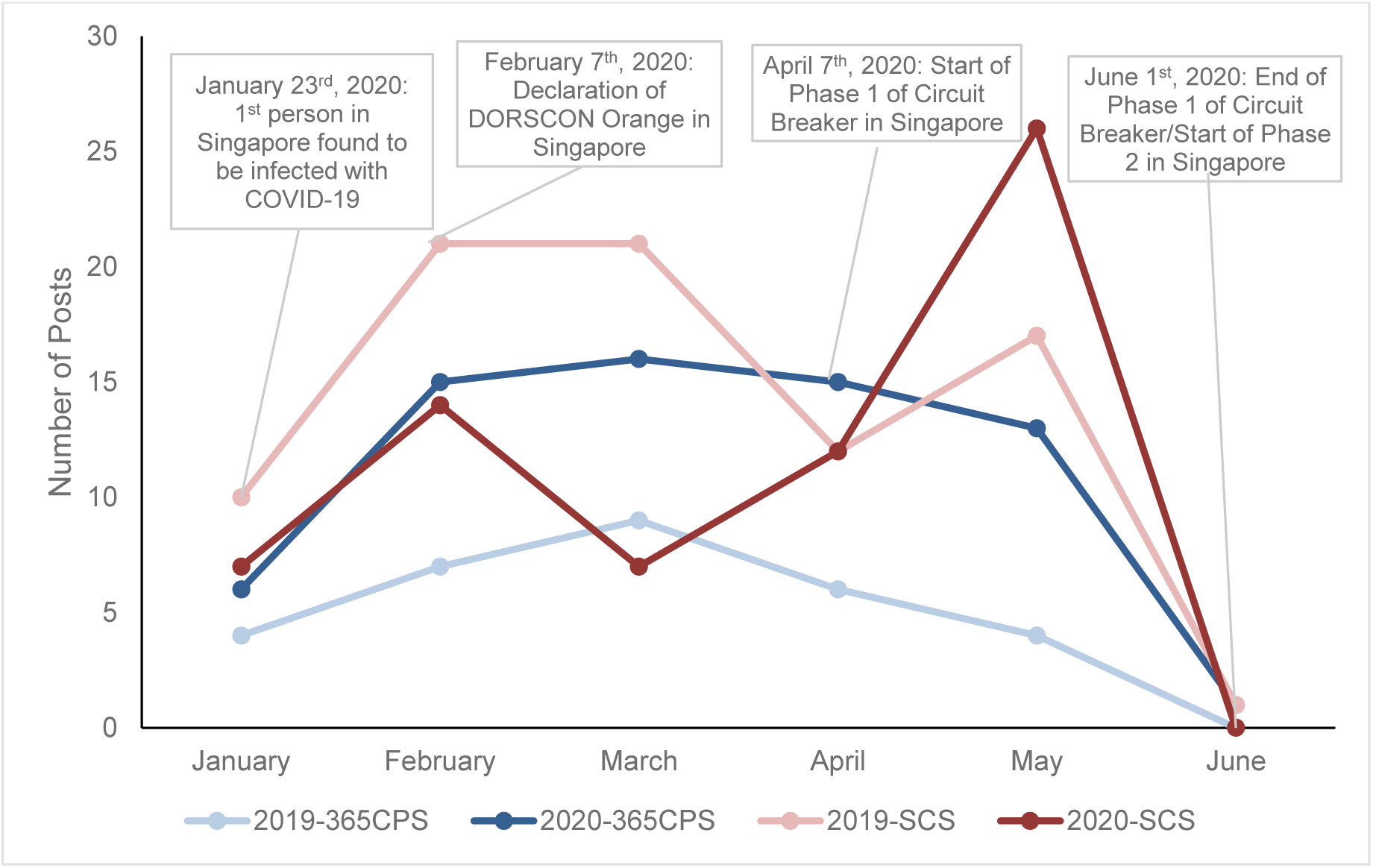
Monthly posts frequency of 365CPS and SCS in 2019 and 2020.

A total of 148 posts were extracted from SCS’s Facebook page and all 148 posts were included in the study. The number of posts was higher in 2019 (n=82) than 2020 (n=66) except for May 2020. SCS started publishing more monthly posts in 2020 since CB started on April 7^th^, 2020. May 2020 also recorded the highest number of monthly posts for SCS (n=26). There was no statistically significant difference between the number of posts by SCS in 2019 and 2020 (z=-1.313, p-value=0.189). However, there was a statistically significant difference when comparing the number of posts by SCS across the three nominated periods in 2020 (z = 2.961, p-value=0.00307).

### 3.2 Identification of Categories for Classifying Facebook Posts

10 categories were identified based on preliminary examination of the Facebook posts extracted from the two cancer-SSAs. In Table 1, the categories and their descriptions are listed. Certain aspects of a cancer survivorship care model proposed in Singapore were also used in conceptualizing the categories [23]. This survivorship care model defines five principles deemed important for survivorship care in Singapore. Two principles were used in the study for developing a few categories. The first principle from the model, ‘Survivor-Centered Care’, was defined as engaging cancer survivors and empowering them to take ownership of their own health and survivorship. This principle was adapted in this study, with categories such as *Educational materials on cancer, Nutritional information and advice*, and *Lifestyle changes*. Posts classified under these categories would be seen as the cancer-SSA’s attempt to provide more information about cancer for the public to educate and empower them. The second principle ‘Promotion of wellness’ was defined to include community-based programs and activities to promote healthy living, self-monitoring and the prevention of the disease itself. This principle was adapted in this study to include categories such as *Promotion of services* and *Promotion of programs and events* [23].

**Table 1.** Categories and their accompanying descriptions

| Category | Description |
| --- | --- |
| Promotion of services | Health and support services organized by SSA or other organizations and supported/promoted by SSA. Services are offered to cancer patients and beneficiaries which include but are not limited to:<br>- Rehabilitation<br>- Transportation to clinic/hospital<br>- Cancer screening activities<br>- Support groups<br>- Day activity centres<br>**Excludes:** Talks hosted/supported by SSAs regarding cancer, exercise activities provided for the general public, E-letters to the public |
| Educational material on cancer | Informative materials provided by SSAs or an invited healthcare expert on:<br>- Cancer symptoms<br>- Cancer Treatment<br>- Risk factors of cancer<br>- Complications of cancer<br>- Side effects of cancer treatment<br>- Cancer prevention and risk reduction<br>- Cancer therapy<br>**Includes:** Provision of external weblinks/opportunity for public to ask more about cancer (such as Q&As), Posted videos for the public to find out more from the SSA/healthcare expert.<br>**Excludes:** Information that is not cancer-related, Educational information provided for the purpose of promoting programs/events/requests for donations |
| Promotion of programs and events | Physical/Online events and activities hosted by SSAs or other organizations and supported/promoted by SSA for their beneficiaries and the public such as but not limited to:<br>- Walkathons or Marathons<br>- World Cancer Day events<br>- Charity events<br>- Online giveaways<br>**Includes:** Posts with signup links for the event/programs, posts describing events/programs that have already been hosted/supported by SSA (such as posts providing a summary of the event), exercise activities shared for the public, volunteering opportunities that may include a signup link, announcement of giveaways, the conducting of the giveaways and the announcement of the winners.<br>**Excludes:** Health and support services provided by SSA (such as health screening) |
| Nutritional information and advice | Information/advice related to food such as:<br>- Healthy eating habits<br>- Consequences of bad eating habits<br>- Healthy recipes<br>- Food safety<br>- Food storage<br>**Includes:** Posts with photos showcasing information listed above (should the post itself have insufficient information provided in the message), links for people to find out more about the types of information listed above, generic nutritional facts that may not be related to cancer<br>**Excludes:** Non-healthy recipes which do not promote healthy living, programs/events inviting beneficiaries to find out more nutritional information, posts which showcase nutritional information for the purpose of inviting the public to attend a program/event related to nutrition |
| Lifestyle changes | Information/advice shared in the post that:<br>- Informs readers on positive lifestyle habits (smoking cessation, weight loss, alcohol consumption, physical/mental exercises) that promotes health in general<br>- Information on consequences of bad lifestyle habits<br>- Promotes lifestyle changes in posts<br>**Includes:** link for people to find out more about lifestyle changes, generic lifestyle habits that may not relate to cancer<br>**Excludes:** Promotion of exercise classes/lessons/events by the SSA for the public, Posts that focuses more on nutritional facts than lifestyle changes, posts that depict past events/programs on lifestyle changes |
| Psychosocial Support | Motivational quotes by people/SSA related to cancer, stories shared by caregiver, volunteer, or cancer patient, pledges by people to support the fight against cancer<br>**Includes:** Online series that share people's stories over a period of time<br>**Excludes:** words of encouragement/psychosocial support provided as a basis to request for donations |
| Donations | Call to the public for donations<br>**Includes:** links to donation sites<br>**Excludes:** Posts that promotes/shares charity events, posts that show appreciation to donors without inviting public to donate |
| Appreciation posts | To Healthcare workers, volunteers, caregivers, organizations that supports/sponsors events with SSAs<br>**Includes:** Description of events where people were allowed to show appreciation to the 3 parties above such as: Writing cards and messages, making items for them, Call to send in appreciation messages and comments |
| COVID-19 related news | Safe distancing practices, information on changes to services and activities due to COVID-19, healthy habits to follow to prevent transmission of disease |
| Others | Posts that do not fall within the above categories |

The category *Psychosocial support* was not adapted from the survivorship care model but was included as cancer is an emotionally taxing disease which affects the patient and their family members. The mental and social health of those affected should be taken into consideration by the cancer-SSAs when publishing posts promoting a positive mindset or encouragement[24]. Other categories such as *Donations* and *COVID-19 related news* were not part of the survivorship care model and were included based on preliminary examination of extracted Facebook posts. Posts that did not fall into any of the abovementioned categories were classified under *Others*.

### 3.3. Cancer-SSAs’ Posts by Category

Table 2 presents the number of posts published by 365CPS per category, to facilitate the comparison between 2019 and 2020, as well as across the three nominated periods in 2020. There are significant differences in the content of the posts between 2019 and 2020. An increase in posts requesting for donations can be seen in 2020 (n=10) compared to 2019 (n=0), as well as posts that promotes educational materials on cancer in 2020 (n=5) compared to 2019 (n=0). Within 2020, there was an increase in the number of posts promoting lifestyle changes from March 2020 to May 2020 compared to January and February 2020. A statistically significant difference was found for the proportion of posts categorized under donations between 2019 and 2020 (χ^2^=5.647, p-value=0.032). There was no statistically significant difference in the number of posts between 2019 and 2020 or across the three nominated periods in 2020 for other categories.

**Table 2.** 365CPS Facebook posts by category

| Category | 2019<br>(n = 30) N<br>(%) | 2020<br>(n = 66)<br>N (%) | P-value | P1 2020<br>(n = 10)<br>N (%) | P2 2020<br>(n = 30)<br>N (%) | P3 2020<br>(n = 26)<br>N (%) | P-value |
| --- | --- | --- | --- | --- | --- | --- | --- |
| Promotion of Services | 3 (10.0) | 2 (3.0) | 0.174 | 0 (0.0) | 1 (3.3) | 1 (3.9) | 1.000 |
| Educational Materials<br>on Cancer | 0 (0.0) | 5 (7.6) | 0.321 | 0 (0.0) | 4 (13.3) | 1 (3.9) | 0.362 |
| Promotion of<br>Programs/Events | 9 (30.0) | 11 (16.7) | 0.176 | 2 (20.0) | 6 (20.0) | 3 (11.5) | 0.665 |
| Nutritional<br>Information/Advice | 12 (40.0) | 16 (24.2) | 0.147 | 4 (40.0) | 6 (20.0) | 6 (23.1) | 0.541 |
| Lifestyle Changes | 5 (16.7) | 8 (12.1) | 0.536 | 0 (0.0) | 2 (6.7) | 6(23.1) | 0.113 |
| Psychosocial Support/Words of Encouragement | 1 (3.3) | 0 (0.0) | 0.312 | 0 (0.0) | 0 (0.0) | 0(0.0) | 0** |
| Donations | 0 (0.0) | 11 (16.7) | 0.016 | 2 (20.0) | 7 (23.3) | 2 (7.7) | 0.241 |
| Appreciation Posts | 0 (0.0) | 2 (3.0) | 1.000 | 0 (0.0) | 1 (3.3) | 1 (3.9) | 1.000 |
| COVID-19 Related News | 0 (0.0) | 5 (7.6) | NA* | 1 (10.0) | 1 (3.3) | 3 (11.5) | 0.449 |
| Others | 0 (0.0) | 6 (9.1) | 0.172 | 1 (10.0) | 2 (6.7) | 3 (11.5) | 0.855 |

*Note: P-values were calculated based on whether there was a statistically significant difference in the proportion of posts under a specific category between 2019 and 2020, and across P1, P2, P3 of 2020. \*P-value was not calculated for the proportion of posts under 'COVID-19 related news' between 2019 and 2020 because COVID-19 only occurred after the timeframe analysed in 2019. \*\*As the number of posts under 'Psychological Support' category is 0 across all 3 periods, the p value cannot be calculated for the test as no difference can be found.*

Table 3 presents the number of SCS posts listed for each category for 2019 and 2020. Similar to 365CPS, there were more posts requesting for donations in 2020 (n=7) compared to 2019 (n=1). More nutritional information was also provided in 2020 (n=4) than 2019 (n=0), especially during CB as can be seen in April 2020 (n=1) and May 2020 (n=3). Between 2019 and 2020, a statistically significant difference was found for the proportions of posts categorized as nutritional information and advice (p-value = 0.038) and for the proportions of posts categorized as donations (p-value=0.022). Across the three periods in 2020, a statistically significant difference was found for the proportion of posts categorized *as* promotions of services (χ^2^=7.99, p-value=0.007) and the proportion of posts categorized as psychosocial support (χ^2^=15.178, p-value<0.0001). The other statistical tests showed no statistically significant difference.

**Table 3.** SCS Facebook posts by category

| Category | 2019<br>(n = 82)<br>N (%) | 2020<br>(n = 66)<br>N (%) | P-value | P1 2020<br>(n = 16)<br>N (%) | P2 2020<br>(n = 14)<br>N (%) | P3 2020<br>(n = 36)<br>N (%) | P-value |
| --- | --- | --- | --- | --- | --- | --- | --- |
| Promotion of Services | 17 (20.7) | 8 (12.1) | 0.190 | 0 (0.0) | 5 (35.7) | 3 (8.3) | 0.07 |
| Educational Materials on Cancer | 6 (7.3) | 4 (6.1) | 1.000 | 0 (0.0) | 1 (7.1) | 3 (8.3) | 0.646 |
| Promotion of Programs/Events | 31 (37.8) | 20 (30.3) | 0.387 | 6 (37.5) | 2 (14.3) | 12 (33.3) | 0.323 |
| Nutritional Information/Advice | 0 (0.0) | 4 (6.1) | 0.038 | 0 (0.0) | 0 (0.0) | 4 (11.1) | 0.318 |
| Lifestyle Changes | 2 (2.4) | 1 (1.5) | 1.000 | 0 (0.0) | 0 (0.0) | 1 (2.8) | 1.000 |
| Psychosocial Support/Words of Encouragement | 10 (12.2) | 6 (9.1) | 0.604 | 6 (37.6) | 0 (0.0) | 0 (0.0) | <0.001 |
| Donations | 1 (1.2) | 7 (10.6) | 0.022 | 1 (6.3) | 0 (0.0) | 6 (16.7) | 0.243 |
| Appreciation Posts | 4 (4.9) | 3 (4.6) | 1.000 | 0 (0.0) | 1 (7.1) | 2 (5.6) | 0.780 |
| COVID-19 Related News | 0 (0.0) | 9 (13.6) | NA* | 3 (18.8) | 3 (21.4) | 3 (8.3) | 0.343 |
| Others | 11 (13.4) | 4 (6.1) | 0.176 | 0 (0.0) | 2 (14.3) | 2 (5.6) | 0.236 |

*Note: P-values were calculated based on whether there was a statistically significant difference in the proportion of posts under a specific category between 2019 and 2020, and across P1, P2, P3 of 2020. \*P-value was not calculated for the proportion of posts under 'COVID-19 related news' between 2019 and 2020 because COVID-19 only occurred after the timeframe analyzed in 2019.*

## 4. Discussion

To our best knowledge, this is the first study which looks at how the COVID-19 pandemic has affected the outreach efforts of cancer-SSAs in Singapore on a social media platform. Compared to 2019, 365CPS published more than twice the number of posts in 2020. This is due to the monthly cancer awareness programs that 365CPS conducted, such as ‘Gallbladder Cancer Awareness Month’ in February 2020 and ‘Colorectal Cancer Awareness Month’ in March 2020 [25]. Perhaps, as physical roadshows cannot be conducted, 365CPS has shifted their focus to promote cancer awareness through Facebook posts. A literature review on social media use by patients concluded that social media platforms such as Facebook provide an excellent tool to rapidly disseminate important health information whilst providing a platform for the public to engage relevant health authorities for further inquiries.[26] Hence, the use of social media by 365CPS will allow them to easily and rapidly disseminate information to their social media followers.

The number of posts by SCS in 2020 was lower than 2019, especially for February and March. There were two events in 2019 which were not repeated in 2020. For example, in February and March 2019, CSC rolled out the ‘I Do’ series and ‘Motivation Monday Posts’ [27,28]. The ‘I Do’ series was a 5-episode series which detailed a love story entangled in cancer. ‘Motivation Mondays’ were a series of encouragement posts published on Mondays to provide emotional support to their followers. We are curious why the event was not repeated or conducted in a modified form in 2020, especially for the ‘Motivation Mondays’ series, which appears to be much needed during COVID-19 pandemic.

Regarding the three nominated periods in 2020, 365CPS posted the most in P2 compared to the other periods. However, the number of posts per day decreases across the three periods (P1: 0.67 posts per day, P2: 0.51 posts per day, P3: 0.48 posts per day). In addition to the cancer awareness months which contributed to an increase in the number of posts from February to April 2020, 365CPS also posted content that encouraged the public to engage in certain health related activities from the comforts of their own homes. This posting pattern coincided with the CB period when the Singaporean government encouraged people to stay home to prevent the spread of the virus [29]. Some of these activities included lymphatic exercises [30] and recipes to boost immunity [31], which allowed beneficiaries to take ownership of their own health during COVID-19 where access to facilities and resources provided by 365CPS were limited.

For SCS, there was an increase in the number of posts in P3 compared to the other periods. On top of regular posts focusing on cancer awareness [32], SCS also informed the public on changes to their services and events due to additional social distancing measures during CB [33,34]. Similar to 365CPS, SCS posted contents related to activities and events which the public could undertake at home during CB. Such content included introducing healthy recipes and activities such as online bingo, a social media trend during CB [35].

While reviewing the contents of the posts according to our classification scheme, we found that the number of posts by SCS promoting services in 2019 was higher than 2020, which could be attributed to the ease of hosting physical events during the pre-COVID period when there was no social distancing measures. Nevertheless, SCS still attempted to reach out to their beneficiaries, encouraging active cancer screening to continue during CB and continuing to provide accessible care for their patients [36].

After an initial quiet period in P1, SCS started to promote more of their services in P2 and P3 of 2020. The promotion of FIT Kits provided by SCS as part of their ‘Colorectal cancer awareness month’ in March 2020 was evident [37]. This also included the promotion of cervical cancer screening as part of their ‘Women’s gynecological cancer awareness campaign’ [38]. It is heartening to see that promotion of cancer prevention is not hampered by CB. However, we are unable to judge the effectiveness of promoting cancer prevention through social media in our study. For example, we do not know if the take-up rates for FIT Kits and cervical cancer screening have increased due to the use of social media platform.

365CPS did not publish any posts related to educational materials on cancer in 2019, but published five posts in 2020 to promote cancer-related information to the public, with most of these posts in P2 of 2020. These posts encouraged regular and early health screening to prevent the risk of cancer [25], and provide more information on the type of psychosocial needs that a cancer patient requires [39]. Whilst the increase in the number of posts in this category from 2019 to 2020, and across the three periods in 2020 was not statistically significant, it can be seen that 365CPS is attempting to provide more resources and information to the public about cancer, encouraging their beneficiaries to take charge of their own health and empowering them to be more invested in their healthcare.

In contrast, SCS had fewer posts on educational materials on cancer in 2020 than 2019. Nevertheless, they started sharing more programs for their followers during P3 compared to the other two periods in 2020. For instance, there was a focus on their interschool competition which encouraged the public to learn more about the projects they had in collaboration with different schools in contributing to their mission of ‘minimising cancer and maximising lives’ [40]. In addition, learning more about the projects provides the public with the benefit of learning more about the topics discussed during the competition, showing how SCS is empowering their followers to continue educating themselves on the different aspects of cancer during COVID-19.

365CPS had more posts requesting for volunteers in 2020. This may be due to a drop in the number of available volunteers to assist beneficiaries during the pandemic for reasons such as social distancing measures and a fear of contracting the virus by being out of the home [41]. Volunteers form a core part of any SSA. Hence, the appeal for volunteers is expected. In addition, 365CPS introduced a series of Lymphatic Detox Exercises that the public could undertake. These were originally physical lessons which had to be suspended during CB [42]. As a result, 365CPS began structuring their exercises such that the public could participate from their homes [43].

We had anticipated an increase in the number of posts related to nutritional information and advice as more people were staying at home and presumably cooking at home. However, we did not observe any statistically significant difference in the number of posts between 2019 and 2020 or across the three nominated periods in 2020. Nonetheless, we do observe a difference in the contents of the posts related to nutritional information and advice. In 2020, there were more posts by 365CPS on healthy recipes whereas in 2019, the contents were related to information on healthy eating and food safety rather than recipes [27]. Providing healthy recipes for their followers to try at home is not the only one way of educating the public in eating healthily to prevent cancer, but also to encourage people to remain at homes and adhere to social distancing measures. SCS did not publish any post related to nutritional information and advice in 2019 but published four such posts in P3 2020. Their posts also share healthy recipes for the public to try at home while during CB, with the hashtag ‘#stayhomeforSG’ indicating that such recipes were meant to encourage people to stay home during CB.

There was an increase in the number of posts related to lifestyle changes for 365CPS from P1 to P3 in 2020. Posts focused on coping with worries and anxieties was a new theme in 2020. These posts reflect the anticipated increase in the level of anxiety among their beneficiaries and other issues that may arise from experiencing either a prolonged confinement in one location, or a change in job situation due to the pandemic [44]. It is encouraging to note that 365CPS is sensitive to the changing needs of their beneficiaries.

We were surprised to note that that the number of posts related to psychosocial support dropped significantly from P1 to P2 and P3 in 2020 for SCS, with no posts from P2 and P3. Most of the posts under this category in P1 was part of SCS’s initiatives during World Cancer Day to showcase local celebrities pledging their support to fight cancer. Psychosocial support is important during the COVID-19 period due to long periods of confinement which may have detrimental effects on one’s mental wellbeing, especially if one has a debilitating condition such as cancer [45]. With CB moving people to more online activities, it is even more important that avenues of support were provided for cancer patients by the SSAs. Unfortunately, this was not the case for both SCS and 365CPS, with the latter having no posts falling under this category in 2020.

In 2020, 365CPS shared more donations related posts. As physical fundraising activities could not be held during CB, SSAs such as 365CPS turned to social media platforms to request for donations. As SSAs rely primarily on donations to sustain their operations, the increase in posts requesting donations from the public is understandable. For 365CPS, their *donations* posts were usually accompanied by more information on how beneficiaries can benefit from the support from the public, and provide links for the public to find out how they can better support beneficiaries [46]. The increase in donations-related posts was also observed for SCS from 2019 to 2020, especially from P1 and P2 to P3 in 2020. For SCS, their request for donations were often tagged with a seasonal greeting on the day of a public holiday, such as Vesak day [47]. An online musical performance was also featured along with a request for donations to SCS [48]. By moving their donation drives online, SSAs may be able to target more age groups cost-effectively, including older internet users, whose numbers have jumped significantly in the past few years [49]. Another benefit of online fundraising is its reach to willing donors who otherwise would not be able to attend physical fundraising activities, thus providing more people with an opportunity to give back to society.

There are certain limitations in our study. Only two cancer-SSAs and their Facebook posts were included in our study. Thus, our findings may not be representative of other cancer-SSAs or other social media platforms. Regardless, Facebook is the most popular social media platform with the highest number of monthly active users. Thus, analyzing Facebook posts would be the most efficient option compared to analyzing other platforms [50]. Another point to note was that the survivorship care model used in this study focused on providing care and support to cancer survivors whilst this study looked at both cancer patients and the general public. Some aspects of the model such as ‘Robust adaptive patient assessment program’ did not apply to cancer-SSAs as their extent of support and care do not including assessing the health issues of cancer patients. Nevertheless, most aspects of the model were applicable in the context of cancer-SSAs providing support and services to their beneficiaries and the general public.

This research can be extended in future studies for analyzing all cancer-SSAs or focus on other social media platforms utilized by these cancer-SSAs such as YouTube or Instagram. Another potential topic for further studies includes analyzing the level of follower engagement achieved through these posts to ascertain whether the cancer-SSAs have successfully achieved an optimum level of engagement with their beneficiaries and the general public.

## 5. Conclusions

Our study aimed to understand the effects of the COVID-19 pandemic on the outreach strategies on Facebook of two cancer-related SSAs for supporting their beneficiaries. We found that 365CPS published more posts in 2020 than 2019, especially during the period where more people were encouraged to stay home. However, SCS published fewer posts in 2020 than 2019, even though in 2020, they published more posts during CB. The different restrictions implemented across the three nominated periods had a slight effect on the type of content published by the SSAs. During P2 and P3, the posts were focused on activities or programs that the public could undertake while staying at home. There was also an obvious increase in the number of requests for donations in 2020, as well as the promotion of lifestyle changes for their beneficiaries to cope with their condition as well as the pandemic. However, posts to provide psychosocial support or encouragement were lacking for both SSAs.

Overall, the efforts taken by the SSAs in adapting to the COVID-19 pandemic and providing important health information and support for their beneficiaries online are applaudable. Our analyses could help SSAs to reflect on their responses to the pandemic. While COVID-19 was unexpected, the same cannot be said of the next pandemic. SSAs should continue learning from the COVID-19 pandemic. One step forward would be to develop a better strategy for their social media posts aimed at not only continuously providing accessible care to the public but also empowering the public to take ownership of their own health, especially during future pandemics.

## Data Availability

Dataset can be provided on request

